# Accelerated approval of non-cancer drugs by the US Food and Drug Administration: A protocol for a meta-epidemiological investigation

**DOI:** 10.1101/2020.05.18.20105148

**Authors:** Kenji Omae, Akira Onishi, Ethan Sahker, Toshi A Furukawa

## Abstract

The US Food and Drug Administration (FDA) ’s accelerated approval program, in which drugs are approved on the basis of surrogate endpoints, has been a source of controversy in the context of risk-benefit trade-offs. The FDA recently reported on the 25-year experience with the program and concluded that the low failure rate in post-approval confirmatory trials was evidence that the program was operating effectively. However, many studies have revealed manufacturers’ poor adherence to obligation of conducting confirmatory trials, and also concerned the harm and cost of cancer drugs approved through this program based on the limited evidence. To date, few studies have investigated accelerated approval of non-cancer drugs. Therefore, the purpose of this study is to review trends in the FDA’s utilization of its accelerated approval program for non-cancer drugs and clarify the status of fulfillment of social obligations by the FDA and sponsors in the program. The FDA’s documents will be reviewed at Drugs@FDA to identify pre-approval and confirmatory trials and regulatory updates for drug indications granted accelerated approval other than cancer treatments. We will further search PubMed, Google/Google Scholar, and the sponsors’ websites to match each identified confirmatory trial to publications in the medical literature.

## INTRODUCTION

For serious or life-threatening diseases, the US Food and Drug Administration (FDA) can approve drugs on the basis of surrogate endpoints that are “reasonably likely to predict clinical benefit,” via its accelerated approval review pathway.^1^ Surrogate endpoints consist of markers, including laboratory values, radiographic images, or other physical measurements, that can reflect true clinical endpoints, such as a reduction in patient symptoms or mortality. In 1992, Congress authorized this accelerated approval program as a response to the demand for faster drug development in the context of the HIV/AIDS crisis.^2^ Since then, the program has expanded to include oncology products and drugs for other diseases, now accounting for about 10% of new drug approvals.^3^

The accelerated pathway commits drug manufacturers to conduct post-approval trials to confirm the drug’s efficacy and safety for the approved indication. Current FDA guidance requires that the confirmatory study be ongoing during initial drug approval. Confirmatory studies must be adequate and well-controlled to monitor both clinical benefit and risk profile. Manufacturers must also include a statement in the official labeling stating clinical benefit “has not been established” and that confirmatory trials are ongoing. The labeling is revised when those confirmatory trials are completed.

The accelerated approval program has been a source of controversy, especially in cancer drug approval. Approving drugs on the basis of surrogate endpoints can be risky, as promising surrogates may later be found not to accurately predict actual changes in patient health outcomes. To make matters worse, it can lead to unnecessary and avoidable adverse events such as detrimental side effects or deaths if it is not properly managed. The FDA recently reported on the 25-year experience with the accelerated pathway. The report evaluated 93 oncology indications granted accelerated approval from December 11, 1992, through May 31, 2017.^4^ The review found that confirmatory trials verified clinical benefit in 51 (55%) of the 93 indications. In 5 cases (5%), approval for an indication was withdrawn based on post-approval trial findings, and for the remaining 37 (40%) indications, post-approval evaluations were ongoing. The FDA concluded that the low failure rate in confirmatory trials was evidence that the accelerated approval pathway was operating effectively. However, many studies have revealed manufacturers’ poor adherence to obligation of conducting post-approval confirmatory studies, and also concerned the harm and cost of cancer drugs approved through this pathway based on the limited evidence.^5-9^ These findings suggest that further efforts and continuous monitoring are needed to properly implement this program in the context of the risk-benefit trade-off.

As mentioned above, accelerated approval of cancer drugs has been intensively studied, while few studies have investigated accelerated approval of non-cancer drugs. Thus, the purpose of this study is to review trends in the FDA’s utilization of its accelerated approval program for non-cancer drugs and clarify the status of fulfillment of social obligations by the FDA and sponsors in the program. We will evaluate the extent to which confirmatory studies were completed, the extent to which accelerated approvals were converted to regular approvals, and the time between accelerated approval and fulfillment of post-approval requirements. The frequency of post-market safety-related label modifications and withdrawals and the publication statuses of completed confirmatory studies will also be examined.

## METHODS

### Sample Identification

We will review publicly available FDA documents (CDER [Center for Drug Evaluation and Research] Drug and Biologic Accelerated Approvals Based on a Surrogate Endpoint) to identify all the indications that received accelerated approval by the FDA since 1992.^10^ We will collect data on all the molecular entities and original therapeutic biologics approved for the indications other than cancer treatment. As with the previous study on cancer drug,^6^ this study will include drug indications approved up to May 2018, resulting in allowing at least 2 years for the completion and publication of post-approval confirmatory studies.

### Identification of Pre-approval Studies

The clinical studies underlying accelerated approval will be identified and characterized. For all drug indications in the sample, the medical review reports and product labels from the Drugs@FDA database will be examined to identify pre-approval studies that established the drug’s efficacy. Drugs@FDA is a publicly available database of all FDA-approved products and contains the approval history for each product, including links to communications from the FDA to the sponsor and product label updates.^11^ When available, medical review reports will be used to gather information about pre-approval trial characteristics. Medical review reports provide a comprehensive overview of the efficacy and safety of a drug. When medical reviews are not available (as can be the case for supplemental approvals), the product labels that describe the key clinical studies supporting the accelerated approval for a new indication will be used.

### Identification of Post-approval Confirmatory Studies

We will review the FDA’s approval letters and medical review reports available in the Drugs@FDA database to identify the post marketing requirements (PMRs), including the schedule of confirmatory studies, at the time of accelerated approval. Information reported in the FDA’s documents will be used to summarize how the FDA characterized the main limitation of the available data at the time of accelerated approval and whether required confirmatory studies have assessed efficacy, safety, or long-term follow-up.

Two sources of information will be reviewed to determine the status of post-approval study requirements. First, we will search the FDA’s publicly available database of PMRs and commitments,^12^ and collect the information on status labels defined by the FDA for post-marketing commitments. Status labels are defined as, (a) “ongoing” – the confirmatory trial is proceeding according to, or ahead of, the original schedule as negotiated between the manufacturer and the FDA, (b) “delayed” – the progression of the confirmatory trial is behind the original schedule, (c) “pending” – the trial has not been initiated and it does not meet the criterion for delayed status, (d) “terminated” – the applicant ended the trial before completion and has not yet submitted a final study report to the FDA, (e) “submitted” – the applicant has concluded or terminated the study, has submitted a final study report to the FDA, but FDA has not yet notified the applicant about commitment fulfillment or release, and (f) “fulfilled” – the FDA is satisfied that the applicant has met the terms of the commitment. Sometimes the FDA will release a manufacturer’s obligation to conduct a post-marketing study because the trial is either no longer feasible or would no longer provide useful information. Such feasibility and utility indications are listed as, (g) “released.”

Second, we will screen ClinicalTrials.gov, a publicly available clinical study registry, for all confirmatory study requirements. Since 2007, Section 801 of the FDA Amendments Act has required the registration of clinical studies subject to FDA regulation, including studies that satisfy post-marketing requirements. For each registered study, the trial registries specify the status (e.g., still recruiting, ongoing but no longer recruiting, completed), as well as the start and end date. The confirmatory study status, complete or ongoing, per specified timelines in the FDA’s approval letters will be assessed. Studies will be considered delayed if the estimated primary completion date in trial registries is at least 1 year later than specified in the FDA approval letters according to the previous study.^8^ When there is a discrepancy between the FDA’s public database and ClinicalTrials.gov, information from ClinicalTrials.gov will be used to determine the status of post-approval study requirements.

Three reviewers (AO, ES and KO) will screen the databases independently. Disagreements will be resolved by discussion; otherwise, a fourth independent reviewer (TAF) will arbitrate.

### Data Extraction in Pre-approval and Confirmatory Studies

Information extracted from each pre-approval and confirmatory study will include design (randomized vs nonrandomized and the status of allocation concealment in a randomized study), comparators, participant enrollment, and primary end point. Comparators will be classified as active (comparing drugs A vs B), add-on (comparing drugs A + B vs drug B alone), placebo, or none. Drugs tested in single-intervention-group trials will be classified as having no comparators. Type of blinding (double blinded vs single blinded vs open label) will be recorded. Study findings will be summarized in terms of the specified primary endpoint. In confirmatory studies, verification of clinical benefit will be assessed.

### Publication Matching of Completed Confirmatory Studies

We will search PubMed, Google/Google Scholar, and the sponsors’ websites to match each identified trial to publications in the medical literature. Studies in all languages will be reviewed as abstracts or full texts. Trials identified in the FDA documents and ClinicalTrials.gov will be matched to publications based on the following characteristics: study identifier (NCT number and/or trial ID), drug name, sample size, dosing schedules, arm number, and primary and secondary outcome measures. Publication matching will be performed independently by three investigators (AO, ES and KO) and disagreements will be resolved by discussion. If consensus cannot be reached, a fourth independent investigator (TAF) will arbitrate.

### Assessment of Regulatory Outcomes

To determine if drug indications granted accelerated approvals later have their labels updated, changes to product labels and the accompanying regulatory letters will be reviewed. The FDA’s correspondence with product sponsors on topics related to the approval and changes in status of their products is publicly available in the Drugs@FDA database. The Drug Safety-related Labeling Changes (SrLC) database provides approved safety-related labeling changes from January 2016 forward.13 Data prior to January 2016 are available on the MedWatch website.^14^

### Statistical Analysis

We will descriptively analyze the trends in the FDA’s utilization of its accelerated approval program for non-cancer drugs over time and determine a timeline of approvals, post-marketing trial results, and the safety-related label update or withdrawal. We will further demonstrate the frequency of termination of confirmatory studies and their publication statuses, the frequency of post-market conversion to regular approvals, and the frequency of safety-related label modifications and withdrawals. Mann-Whitney U test and chi-square test will be used to evaluate differences in study features between pre-approval and confirmatory studies, including enrollment, study design, and primary endpoints. A P-value of <0.05 will be considered statistically significant. Statistical analyses will be conducted using STATA version 16 (Stata Corp LP, College Station, TX, USA).

### Ethical Consideration

The protocol for this meta-epidemiological investigation was registered with the University Hospital Medical Information Network (www.umin.ac.jp/ctr/index-j.htm; registration number UMIN000039281). This study will not involve individual patient information; it will involve publicly available trial-level data, and therefore, institutional review board approval will not be required.

## Data Availability

All data generated during the current study will be available from the corresponding author on reasonable request.

## Notes

### Competing Interest Statement

KO reports a speaker fee from Ono Pharmaceutical and Astellas Pharma, and a consultant fee from Kyowa Hakko Kirin, outside the submitted work. AO reports a speaker fee from Chugai, Ono Pharmaceutical, Eli Lilly, Mitsubishi-Tanabe, Asahi-Kasei, and Takeda, and research grant from Advantest, outside the submitted work. TAF reports personal fees from Mitsubishi-Tanabe, MSD and Shionogi, and a grant from Mitsubishi-Tanabe, outside the submitted work; TAF has a patent 2018-177688 pending. ES declares that there is no competing interest.

### Clinical Trial

UMIN000039281

### Funding Statement

This study does not receive any specific grant from funding agencies in the public, commercial or not-for-profit.

## REFERENCES

1. Guidance for Industry Expedited Programs for Serious Conditions – Drugs and Biologics https://www.fda.gov/media/86377/download Accessed October 7, 2019.

2. Johnson JR, Ning YM, Farrell A, Justice R, Keegan P, Pazdur R. Accelerated approval of oncology products: the Food and Drug Administration experience. J Natl Cancer Inst 2011; 103: 636–44.

3. Kesselheim AS, Wang B, Franklin JM, Darrow JJ. Trends in utilization of FDA expedited drug development and approval programs, 1987–2014: cohort study. BMJ 2015; 351: h4633.

4. Beaver JA, Howie LJ, Pelosof L, et al. A 25-year experience of US Food and Drug Administration accelerated approval of malignant hematology and oncology drugs and biologics: a review. JAMA Oncol. 2018;4(6):849–856.

5. Gill J, Prasad V. A reality check of the accelerated approval of immune-checkpoint inhibitors. Nat Rev Clin Oncol. 2019. doi: 10.1038/s41571-019-0260-y.

6. Gyawali B, Hey SP, Kesselheim AS. Assessment of the Clinical Benefit of Cancer Drugs Receiving Accelerated Approval. JAMA Intern Med. 2019; 179(7): 906–913.

7. DiMagno SSP, Glickman A, Emanuel EJ. Accelerated Approval of Cancer Drugs-Righting the Ship of the US Food and Drug Administration. JAMA Intern Med. 2019;179(7):922–923.

8. Naci H, Smalley KR, Kesselheim AS. Characteristics of Preapproval and Postapproval Studies for Drugs Granted Accelerated Approval by the US Food and Drug Administration. JAMA. 2017;318(7):626–636.

9. Gellad WF, Kesselheim AS. Accelerated Approval and Expensive Drugs – A Challenging Combination. N Engl J Med. 2017;376(21):2001–2004.

10. CDER Drug and Biologic Accelerated Approvals Based on a Surrogate Endpoint https://www.fda.gov/drugs/nda-and-bla-approvals/accelerated-approvals Accessed May 11, 2020.

11. Drugs@FDA: FDA Approved Drug Products https://www.accessdata.fda.gov/scripts/cder/daf/ Accessed May 11, 2020.

12. Postmarket Requirements and Commitments https://www.accessdata.fda.gov/scripts/cder/pmc/index.cfm Accessed May 11, 2020.

13. Drug Safety-related Labeling Changes (SrLC) https://www.accessdata.fda.gov/scripts/cder/safetylabelingchanges/ Accesses May 11, 2020

14. MedWatch Medical Product Safety Information archive http://wayback.archive-it.org/7993/20170110235327/http://www.fda.gov/Safety/MedWatch/SafetyInformation/default.htm Accessed May 11, 2020

